# How many lives do COVID vaccines save? Evidence from Israel

**DOI:** 10.1101/2021.10.27.21265591

**Authors:** Ronen Arbel, Candace Makeda Moore, Ruslan Sergienko, Joseph Pliskin

## Abstract

**Background:** In December 2020, Israel began a mass vaccination program with the rapid rollout of the Pfizer-BioNTech COVID-19 BNT162b2 vaccine for adults in Israel. The campaign vaccinated fewer people than necessary for herd immunity. However, at the same time, government stringency measures in terms of closing public life were decreased. Real-world observational data were used to examine the effect of mass vaccination on Covid-19 mortality.

**Methods:** The study period to examine the effect of vaccination on mortality was chosen to capture when at least 90% of the population over age 70 were vaccinated for less than seven months. Projected deaths as expected from vaccine efficacy and actual mortality data were compared for the study population with examination of potential confounding effects of government stringency. Average government stringency (Oxford Stringency Index) was calculated in the study period and the preceding period of the pandemic. Potential confounding effects of an age shift in the distribution of deaths were examined by analyzing the distributions of deaths and cases before and after the study period.

**Results:** Confirmed deaths from COVID-19 in the population over 70 after mass vaccination were recorded as 370, versus 408 expected from applying person-days of vaccine efficacy, and 5,120 estimated without vaccinations.

**Conclusions:** Vaccines against COVID-19 saved more lives than expected by simply applying individual vaccine efficacy to the vaccinated population in Israel, despite a loosening of government stringency.

## INTRODUCTION

In December 2020, Israeli health services managed one of the fastest rollouts of COVID-19 vaccinations of any country (1). Herd immunity, typically estimated by 1-1/R0 (2), requires enough vaccination and/or natural immunity to drive a virus’ reproductive number below 1 over a sustained period. Levels of total vaccination that would induce herd immunity were not reached in Israel (5), and several more transmissible strains entered the country during the vaccination campaign.

In Israel, the Pfizer-BioNTech COVID-19 BNT162b2 was proven efficacious on the level of individuals (3), but the vaccine campaign initially did not target young people, nor was the uptake among different ethnoreligious groups (2). This uneven uptake created a theoretical possibility of a repository for evermore lethal disease to spread (4) (5). However, some researchers posited that vaccinating older people would positively affect the total population level (6). In theory, easing the burden on hospitals might reduce deaths as increased numbers of hospitalized patients had been shown to have led to higher excess mortality (7).

The question of whether vaccinating most older people would achieve dramatic reductions in transmission and mortality or simply push transmissions and deaths into lower-age populations did not have a clear empirical answer. Potential real-world effects of a mass vaccination campaign against COVID-19 such as in Israel were unknown. Therefore, we set out to examine the real-world population-level effects of the vaccination campaign and compared them to models based on proven vaccine efficacy.

## METHODS

### Primary outcomes

The primary outcome was the number of deaths saved by the mass vaccination campaign in the population over 70 in the short term. Thus, the primary outcome includes actual deaths and deaths as predicted by vaccine efficacy for this group during the study period.

### The study period for primary outcome

We examined deaths during the period when at least 90% of the most vulnerable population age group, the population over 70, was vaccinated. Deaths were tallied per age group until the Delta variant began to cause a new wave of infections. Thus we computed deaths by summing deaths in the 104 days from March 15, 2021, until June 26 (included).

### Modeling the effect of vaccinations on mortality

The expected and confirmed deaths in the population by age group were derived from tabular data provided by the Israel Ministry of Health for ten-year cohorts (8).

The effectiveness of vaccinations was estimated by published data on the Israeli population (3). Projected deaths with vaccination were derived by applying the reported deaths for vaccinated participants per person-days from the study to the vaccinated group’s person-days from March 14, 2021, until June 26, 2021. In order to account for slight changes in the percentage of each group vaccinated during this period, sums were made on a weekly basis. Projected deaths without any vaccination campaign were derived by applying the recorded deaths per unvaccinated person-days in the study over the number of person-days to all relevant populations. The confirmed deaths and projected deaths with and without vaccination in the population over 70 are reported.

### Secondary outcomes

To minimize potential confounding effects, case and death distributions were compared in the study period with case and death distributions before the study period for the primary outcome.

To evaluate the potential Covid-19 deaths saved, potential death counts by statistical regression to polynomial functions were created through curve fitting.

As death rates in specific groups might be confounded by changing government policy, we derived and compared average government stringency as quantified by the Oxford Stringency Index (OSI) for the study period for the primary outcome and the period before the study period for the primary outcome. We also calculated the days in total lockdown, defined by an OSI of 80, during and before the study period.

### Computational methods and availability

All calculations were made in Python 3.8 and are available as coded in Jupyter notebooks on the laboratory Github (9).

## RESULTS

### Primary outcomes

Recorded deaths from COVID-19 between March 14 and June 26, 2021, in the population over 70 years old were 370. In this period, there were 930 COVID-19 deaths in the total population (all ages). Given the vaccination rate, the estimated number of deaths in the Israeli population over 70 based on individual vaccine efficacy was 408. Without any vaccines, all other things equal, including the lowered government stringency, the expected number of deaths in the period would have been 5,120.

### Secondary outcomes

The confirmed number of cases in the population over 70 during the study period was 1,119. The death and case distributions were essentially unchanged in their trends before and during the study period. The two distributions for deaths by age before and after the vaccination campaign were 94.08% similar after normalization based on mean squared error. The two distributions for cases were 98.65% similar after normalization based on mean squared error. The skew and kurtosis of cases before and after the study period was very similar. Before and during the study period, the skewness of cases was positive, with more cases in the young. The kurtosis of cases was 7.03 before the study period and 8.38 afterward. The kurtosis of the distribution of deaths before the study period was 6.27, and during the study period was negative 0.28, but the trend towards more deaths increasing with age was maintained.

These distributions of deaths and cases are displayed below in Figure 1:

**Figure 1:**
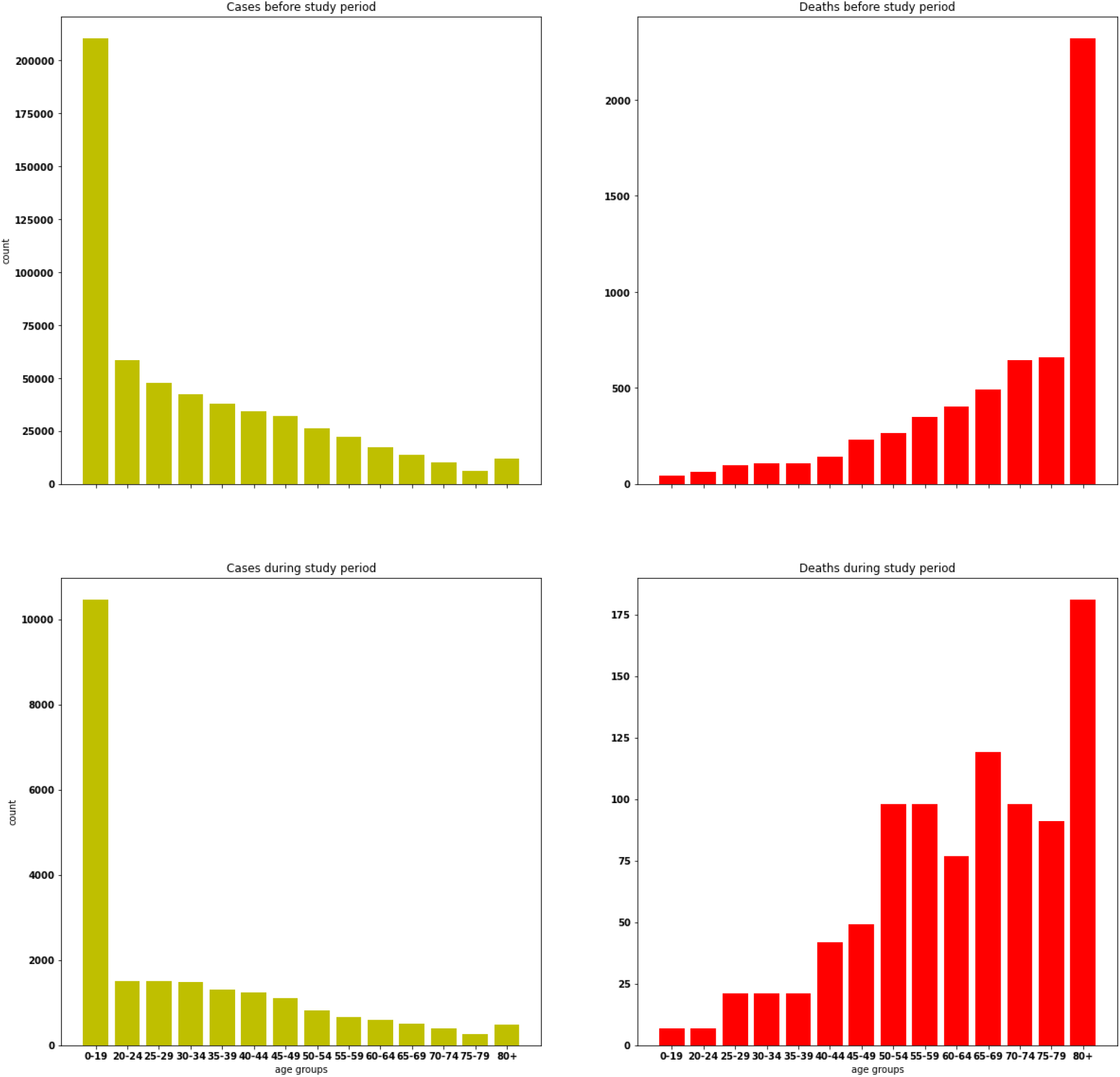
Cases and deaths, before and during the study period

Stringency index before and during the study period was different. Before the study period, Israel’s OSI averaged 61, a maximum of 94, and 128/438 days over 80 (128 days of national lockdowns). However, the OSI during the study period was much lower – averaging 50 with a maximum of 60, and there were zero days with OSI over 80 (no lockdowns).

## DISCUSSION

### Summary of Results

After vaccines were introduced and had significant uptake, government stringency was lower than in previous periods. Once a level of 90% vaccination was reached in the older population, the distribution of cases and deaths was similar across age groups as before, but fewer than expected deaths were seen even as government stringency fell. These results raise the possibility of a pseudo-herd immunity type effect.

### Limitations

Our study was limited in that it focused on the population of individuals over 70 years old. The number of deaths in this age group was 61% of total deaths before mass vaccination of the older population, i.e., before the study period. During the study period the percentage of deaths in this group was 40%.

The study period after vaccinations was limited due to uncertainties related to the Delta variant and the temporal period of vaccine efficacy in individuals. Therefore we limited the study period to the end of June before the Delta variant created new patterns of rising cases. Thus, the precise results relate to conditions in which the Alpha variant was predominant, and the vaccine efficacy was presumed to be high. Many questions about the effects of vaccination on the Delta variant remain unanswered with real world data as it is presently in clear how much of the rise in deaths may be due to vaccines wearing off.

Calculating the precise number of saved lives by any vaccine is an impossible task. The counterfactual number of deaths if the public had never been vaccinated depends upon many variables that change because of the patient’s vaccine uptake. For example, we might assume that due to the high uptake of the COVID-19 vaccine in Israel, the government became less strict in terms of closures, but how strict exactly it might have been in the counterfactual world is a mystery and only one of many variables affecting transmissions and deaths. After a steep rise in deaths, we expect the government to become very strict and perhaps employ novel measures and increasing surveillance to lowering the deaths, so the estimate of deaths without any vaccinations may be overestimated.

Another reason deaths without vaccinations may be overestimated is that there is statistical confounding related to the population’s characteristics that do not vaccinate despite freely available vaccinations and universal health coverage. This small percentage of the population may be generally non-compliant or specifically somehow different in compliance with measures against COVID-19 transmission, and therefore have a different death rate.

Despite the imprecision of our results regarding predicted deaths without vaccination, they were lower than predicted deaths from the results of simple curve-fitting to model the situation with no vaccines and no increasing government stringency, which implies they are within the range of plausibility (10).

Our estimates may help examine whether vaccine efficacy on a population level after a mass vaccination campaign can be predicted from individual-level vaccine efficacy.

Our lack of long-term experience with COVID-19 vaccines makes it unclear how much immunity against future variants they will provide over time. Initial evidence suggests that there is a lessening of vaccine efficacy over time (11). However, we cannot predict how many lives will be saved over many years as new, very varied variants may emerge.

### Strengths

Our research demonstrates that there are empirically additional lives saved over that expected by vaccine efficacy with a strategy of targeting the adult population even when confounders of government stringency and case distribution in age groups are accounted for. We have demonstrated that the real-world effects of a vaccination campaign are not necessarily simply the sum of expected efficacy multiplied over a population, but instead, there is some synergistic effect similar to herd immunity.

## Conclusions

Vaccination in Israel proved to be a better-than-expected tool against the COVID-19 virus’ early variants. There appear to be additional beneficial effects in terms of lives saved of a vaccination campaign in the elderly over what would be expected by applying expected efficacy.

## Data Availability

This study uses raw data obtained from the Israel Ministry of Health and affiliated government websites. All calculations on this data were made in Python 3.8 and are available as coded in publicly available Jupyter notebooks on the laboratory Github (9).

https://github.com/maxcorlabs/Corona_Cost_Effectiveness

## Notes

### Competing Interest Statement

The authors have declared no competing interest.

### Funding Statement

This study was funded by the Israeli National Institute of Health Policy.

### Author Declarations

This study involves only openly available human data on a population level, which can be obtained from the Israel Ministry of Health and affiliated government websites.

